# Development and implementation of pharmacy PrEP awareness raising and initiation: acceptability and feasibility of a UK pilot

**DOI:** 10.1101/2025.07.29.25332355

**Authors:** China R. Harrison, Lindsey Harryman, Sarah Stockwell, Hannah Family, Joanna Kesten, Sarah Denford, Jennifer Scott, Caroline Sabin, Joanna Copping, John Saunders, Ross Hamilton-Shaw, Natalie Symonds, Osarenoma B. Dick, Emma Tarmey, Jeremy Horwood

**Affiliations:** National Institute for Health and Care Research Applied Research Collaboration West (NIHR ARC West) at university hospitals Bristol and Weston NHS foundation Trust, Bristol, UK; National Institute for Health and Care Research, Health Protection Research Unit (HPRU) in Evaluation and Behavioural Science. Population Health Sciences, Bristol Medical School, University of Bristol, Oakfield House, Oakfield Grove, BS8 2BN; Population Health Sciences, Bristol Medical School, University of Bristol, Canynge Hall, 39 Whatley Road, Bristol, BS8 2PS, UK; University Hospitals Bristol and Weston NHS Foundation Trust, Bristol, BS1 3NU; Department of Sexual Health, Cardiff Royal Infirmary, Cardiff & Vale University Health Board. Cardiff, CF24 0SZ; Institute for Global Health, UCL, Royal Free Campus, Rowland Hill Street, London, NW3 2PF UK; NIHR HPRU in Blood-Borne and Sexually Transmitted Infections at UCL, London, UK; Communities and Public Health, Bristol City Council, College Green Bristol BS1 5TR; Blood Safety, Hepatitis, STI & HIV Division, UK Health Security Agency, 61 Colindale Avenue, London, NW9 5EQ, UK; Gilead Sciences Ltd. 280 High Holborn, London WC1V 7EEL; Public Health and Wellbeing, South Gloucestershire Council, South Gloucestershire, BS37 5AF; Patient and public involvement contributor, Bristol, UK

**Keywords:** PrEP, HIV, Community pharmacy, pilot, community members, awareness raising, PrEP initiation, COM-B, HIV prevention

## Abstract

**Objectives:** To develop and implement a community pharmacy pre-exposure prophylaxis (PrEP) awareness raising and initiation pilot, informed by a literature review and interviews with pharmacists and community members.

**Methods:** Using the Person-Based Approach and the Capability, Opportunity, Motivation– Behaviour change (COM-B) model, the pilot aimed to improve HIV/PrEP knowledge/capability and motivation through pharmacist and community awareness-raising. Opportunity was addressed by integrating PrEP consultations into pharmacy services and offering STI, HIV, and estimated glomerular filtration rate (eGFR) home test kits. Financial incentives also aimed to motivate pharmacists. Pharmacists identified eligible community members, initiated PrEP discussions, and provided home testing kits. Individuals opting to start PrEP completed the tests and posted them to a local sexual health clinic, where staff reviewed the results, confirmed eligibility, and prescribed PrEP remotely. NHS policy restrictions on pharmacies stocking NHS-procured PrEP informed the delivery model. The pilot evaluation focused on staff training impact, number/type of consultations, and feasibility and acceptability of pharmacy PrEP delivery for staff and community members.

**Results:** The pilot was conducted in five pharmacies across Bristol, North Somerset, and South Gloucestershire from October 2024 to April 2025. A total of 55 consultations were held with individuals aged 18–69 (n=23 male, n=31 female, n=1 transgender). Visit reasons included emergency contraception (n=14), PrEP (n=14), and opioid substitution therapy (n=11). Twenty-eight community members expressed interest in PrEP; 22 accepted a home testing kit, seven returned it, and four were prescribed PrEP. Pharmacists and community members viewed the service positively. Barriers included pharmacists’ initial lack of confidence initiating PrEP discussions and limited public awareness of PrEP availability in pharmacies.

**Conclusion:** Community pharmacies represent a promising site for PrEP delivery. Successful scale-up requires pharmacist training, public health education, structural and policy changes to support accessible PrEP provision beyond sexual health clinics to reduce inequities.

**Key messages:** *What is already known on this topic:* There are inequities in access to and uptake of PrEP due to a lack of awareness of PrEP, stigma, geographical proximity and access to sexual health clinics. PrEP awareness raising and initiation via community pharmacies could improve access and uptake.

*What this study adds:* This is the first UK pharmacy-based PrEP awareness raising and initiation pilot targeting underserved community members not currently accessing PrEP. Findings show pharmacy PrEP awareness raising and initiation to be acceptable and feasible for pharmacists and community members. Pharmacies have the potential to be valuable community assets for PrEP in addition to traditional sexual health clinics, particularly for underserved community members. Improved community awareness of PrEP and pharmacist confidence in delivering the service could help optimise pharmacy PrEP delivery.

*How this study might affect research, practice or policy:* Community pharmacy PrEP delivery needs to be supported by appropriate training that equips pharmacists with the capabilities and confidence required to provide a comprehensive service. This should be facilitated by policy frameworks and funding that allow provision of PrEP outside of sexual health clinics.

## Introduction

Pre-exposure Prophylaxis (PrEP) is a highly effective HIV prevention method, significantly reducing the risk of HIV acquisition [1]. Since its introduction, PrEP has played a critical role in reducing new HIV acquisitions globally, particularly among gay, bisexual and other men who have sex with men (GBMSM).

In the United Kingdom, PrEP is only currently available free of charge from publicly funded sexual health services. Those initiating PrEP undergo baseline and regular follow up screening tests for sexually transmitted infections (STIs), HIV and kidney function [2]. Consequently, in the UK PrEP is currently primarily accessed via face-to-face clinic-based services and overseen by sexual health clinicians. The clinic-based model of PrEP provision has raised concerns that it may be limiting access for certain populations among whom HIV acquisitions remain high but PrEP use low, for example, Black African and Black Caribbean heritage communities, people who inject drugs (PWID) heterosexual and bisexual women and transgender people, meaning that PrEP remains underused, contributing to persistent health inequalities [1, 3, 4].

Previous research has highlighted barriers to PrEP access, including the geographical distance of sexual health clinics, limited appointment availability and long wait times [5-7], and stigma associated with attending sexual health clinics, particularly for individuals from communities with cultural taboos surrounding sexual health [8-10]. These barriers underscore the need to explore alternative models of PrEP initiation that can enhance the accessibility and uptake of PrEP among underserved populations and reduce inequalities in access.

Recognising these challenges, the UK government, national PrEP guidelines and various research studies have proposed the expansion of PrEP provision to community pharmacies and other settings [11-14]. Pharmacies offer several advantages that could facilitate PrEP uptake.

Pharmacies are widely distributed across urban and rural areas and have extended working hours, providing greater and more convenient accessibility for individuals who may face difficulties accessing sexual health clinics. They also offer a wide range of healthcare services and potentially allow individuals to access PrEP easily within a setting that may feel less stigmatising than a dedicated sexual health clinic [7, 15]. Pharmacists are also experienced in providing health information, so could ensure that individuals interested in initiating PrEP receive comprehensive consultations and care.

Numerous international studies have demonstrated the effectiveness of pharmacy-based PrEP delivery for increasing PrEP awareness and access to underserved populations [16, 17]. Our previous research on the barriers and facilitators of community pharmacy PrEP delivery also highlighted the acceptability of pharmacy-based PrEP initiation [7, 15]. This study aimed to develop, implement and examine the acceptability and feasibility of a UK pharmacy-based PrEP awareness raising and initiation pilot (hereafter pilot), targeting underserved community members not currently accessing PrEP.

## Methods and materials

A mixed methods design was used to develop and evaluate the pharmacy PrEP delivery intervention.

### Pharmacy PrEP Intervention Development

The pilot design was guided by the Person Based Approach (PBA) [18]. The PBA is an iterative approach to intervention development, implementation and evaluation combining evidence, stakeholder and patient and public involvement (PPI), behaviour change theory and mixed methods research. The first stage involved identifying the behavioural facilitators and barriers to pharmacy PrEP delivery for community members and pharmacists using the Capability, Opportunity, Motivation, Behaviour (COM-B) model of behaviour change [19]. COM-B posits that a behaviour (B), e.g. PrEP initiation, is influenced by an individual having (1) the capabilities (C) (physically (skills) and psychologically (knowledge)), (2) opportunity (O) (social (societal influence) and physical (environmental resources) and (3) motivation (M) (automatic (emotion) and reflective (beliefs, intentions)). Informed by COM-B we conducted a scoping review of the literature and qualitative interviews with pharmacists and community members from underserved communities [7, 15].

The second stage involved coproducing the pilot design and accompanying promotional materials with community members to ensure its design met the needs of those who would benefit from PrEP and are less likely to access the sexual health clinic for it. We conducted three workshops with stakeholders (pharmacists, sexual health clinicians, local authority commissioners) and two online patient and public involvement (PPI) meetings with four PPI members.

Results from the scoping review [15] and interviews [7] were collated with the feedback from stakeholders and PPI members (both positive and negative) to create the guiding principles (intervention design objectives and intervention features that achieve each objective) and plotted in an intervention planning table. This iterative process pointed to the need for the intervention to enhance capabilities and motivation through the provision of staff training and community member awareness raising and education via advertising and online information. Additional strategies included increasing opportunity by integrating PrEP discussions into specific consultations and pre-existing pharmacy services and offering home STI (gonorrhoea, Chlamydia, syphilis, Hepatitis B and C, HIV and kidney function (eGFR) screening test kits. The provision of home screening test kits mitigated the need for pharmacists to deliver sensitive test results and manage system linkage requirements with sexual health services. Because community pharmacists were unable to procure PrEP at affordable prices, the pilot involved collaboration with the sexual health clinic. This enabled people to be prescribed PrEP via the sexual health clinic. To increase staff motivation, a £30 financial remuneration (in line with similar services) was provided to pharmacies for each PrEP consultation regardless of whether the community member went on to initiate PrEP.

### Pharmacy PrEP Intervention Pilot Design

All staff working in the pilot pharmacies received online HIV/PrEP educational training adapted from Terrence Higgins Trust training [20]. Pharmacists in each pilot pharmacy received two additional in-person training sessions, delivered by the research team and sexual health clinicians. These training sessions were designed to answer any questions they had, provide them with further PrEP knowledge and support pharmacists to initiate PrEP discussions by, enabling them to share their learnings with each other, and practising discussing PrEP in different consultation situations (e.g., emergency contraception, opioid substitution). The first in-person session was delivered prior to pilot implementation and the second three months into the pilot (December 2024). The first author (CH) regularly visited the pilot pharmacies to check in with the pharmacists and answer any questions they had.

To raise awareness of the pilot among key community populations, posters were distributed in the pilot pharmacies, local community spaces (e.g., community centres, notice boards, doctors’ surgeries), and via community organisations (e.g., Bristol Drugs Project, Refugee Women of Bristol). Digital posters were also distributed online via community organisations sharing on social media and WhatsApp groups. Paid social media advertising on Facebook and Instagram ran from the 14^th^ -23^rd^ February and 3^rd^-4^th^ of March 2025. Pilot promotional material directed individuals to the five pharmacies and the pilot website with information on HIV, PrEP and details of the pilot.

The intervention consisted of seven steps:

1. Community members either i) attend the pharmacy and ask for a PrEP consultation or ii) attend the pharmacy for another reason and the pharmacists introduce PrEP.
2. A PrEP consultation takes place in a private consultation room with a pharmacist.
3. The community member decides if they are interested in starting PrEP.
4. If they are interested, the pharmacist provides the community member with a home screening test kit and an electronic referral is automatically sent to the sexual health clinic.
5. The community member completes the screening test kit at home and using the pre-paid packaging, sends it to the local sexual health clinic for processing.
6. The sexual health clinic phone the community member to discuss their test results, and PrEP eligibility.
7. If PrEP is agreed, the sexual health clinic prescribe and post the PrEP to the community member’s home.

Pharmacists were guided through the PrEP consultation by completing PharmOutcomes® (a secure computer clinical service platform used by community pharmacies). The automatic referral sent to the sexual health clinic via PharmOutcomes® (step 4), notified the sexual health clinic of contact details (name, phone number) of the individuals taking a test to enable the clinic to contact the individual to discuss their test results, PrEP and organise posting PrEP.

### Study Setting

The pilot was conducted in five pharmacies in Bristol (n=3), North Somerset (n=1) and South Gloucestershire (n=1) (BNSSG) between October 2024 to April 2025. Pharmacies in this area were provided with study information by the Local Pharmaceutical Committee. Pharmacies that returned expressions of interest were selected to include maximum variation in relation to HIV prevalence areas, ethnic diversity of the local community, rural or urban location and number of sexual health related consultations provided.

### Study Population

Community members were eligible to participate in the pilot if they were 1) ≥18 years, 2) not currently using PrEP, 3) HIV-negative and 4) able to consent to participate in the study. All pharmacy staff working in the pharmacies were eligible to participate in the online training. Pharmacists were eligible to participate in the interviews if they were involved in the delivery of PrEP consultations.

### Quantitative evaluation methods Training Evaluation

To examine the impact of training, all pharmacy staff receiving online training were asked 10 questions assessing knowledge about HIV and PrEP before and after completing the training [20]. Participants were asked to rate statements as true, false or don’t know. Correct answers were assigned one point and an incorrect score or don’t know answer zero points.

### Pilot consultations and initiations

The number and type of PrEP consultations, in addition to number of test kits dispensed were recorded via PharmOutcomes®. The PrEP PharmOutcomes® template was designed specifically for the pilot so that for each consultation, pharmacists could record the participants age, gender, postcode, ethnicity, phone number, reason for attending the pharmacy, interest in starting PrEP, whether a home testing kit was provided, and if not why. PharmOutcomes® also enabled an electronic referral to be sent to the sexual health clinic if the participant was interested in starting PrEP.

Number of test kits returned, and PrEP prescriptions initiated were recorded by the local sexual health clinic. For each kit returned and PrEP prescription initiated, the sexual health clinic also recorded the participants age, gender, ethnicity, and whether they were known to the sexual health clinic already.

### Qualitative evaluation

Longitudinal interviews were conducted with the pharmacists at 3 and 6 months to explore implementation processes and changes in the acceptability and feasibility of pharmacy PrEP delivery across time. Pharmacists were invited to an interview via email by CH and provided with a participant information sheet. Pharmacists were offered a £40 shopping voucher for their time.

Community members were invited by the pharmacists delivering the consultations to complete an online or paper survey regarding their views and experiences of the PrEP consultation. At the end of the survey, participants were invited to provide their email address to receive details about participating in an interview. An additional text message inviting community members to complete the survey was sent out by the sexual health clinic to those referred for PrEP who had not already completed the survey, all participants who expressed an interest in being interviewed were contacted by CH with a participant information sheet and invited to arrange a time to be interviewed. Anyone who did not respond to the invitation was sent two reminders.

Community members were offered a £20 shopping voucher for their time.

### Data analysis

Quantitative data was analysed using SPSS. A Wilcoxon signed-rank test was used to measure any significant difference between pre and post knowledge scores. Descriptive statistics (means (M), standard deviations (SD) and percentage)) were used to analyse the data provided by the pharmacies and the sexual health clinic concerning the number of PrEP consultations and demographics of community members.

Interviews lasting between 30-45 minutes were conducted online, by telephone or in person by CH using flexible topic guides to explore the views and experiences of pharmacy PrEP delivery. Interviews were audio recorded, transcribed, imported into NVivo software and analysed thematically [21]. Firstly, the transcripts were read several times, to gain familiarisation with the data and initial ideas noted. The transcripts were then examined on a line-by-line basis with inductive and deductive codes being assigned to the segments of the data that provided insight into the participants’ views and understanding of their experiences (i.e., the aims of the process evaluation). Qualitative data for the community members and pharmacists were analysed separately and concurrently, before being triangulated with the quantitative data to provide a more nuanced understanding. Data was coded by the first author (CH, a behavioural scientist and sexual and reproductive health researcher) and reviewed by HF (a psychologist and pharmacy researcher) to sense check interpretation and provide an alternative perspective on the data. Themes were generated by reviewing and grouping connecting codes. The coding and themes were discussed by the multidisciplinary research team to ensure transparency, coherence and commitment to rigor.

## Results

### Evaluation

#### a. Staff training evaluation

All 31 customer facing staff working in the five pharmacies completed the online training. A Wilcoxon signed rank test indicated a statistically significant difference from pre (M=4.77, SD=3.03) to post (M=9.10, SD=1.90) training knowledge scores, (Z=-4.47), *p*<.001) (see figure 1). Ten pharmacists working at the five pharmacies completed additional in-person training.

**Figure 1.**
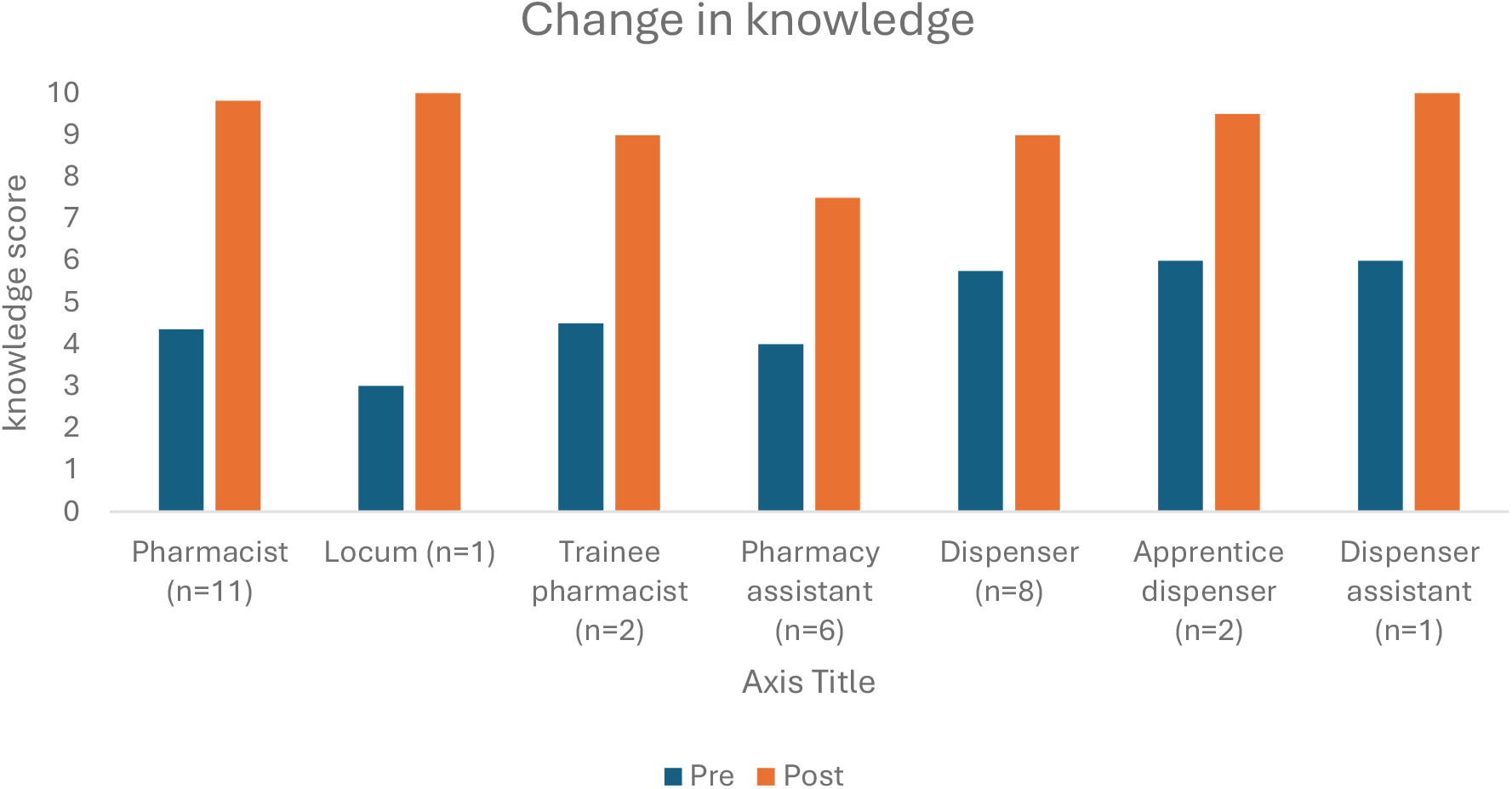
Pre and post online training for the different customer facing staff in the five pilot pharmacies.

#### b. Pilot consultations and initiations

A total of 55 community members had a PrEP consultation between October 2024 and April 2025. Most consultations were with individuals aged 20-29 years, who identified as female and white British (See Table 1).

**Table 1.** demographic characteristics of community members (N=55) having a community pharmacy PrEP consultation, taking a test kit, returning a test kit and being prescribed PrEP.

|  | Pharmacy PrEP consultation (N=55) | Test kit received (n=22) | Test kit returned (n=9) | Prescribed PrEP (n=4) |
| --- | --- | --- | --- | --- |
| **Characteristic** |  |  |  |  |
| **Gender** |  |  |  |  |
| Male | 23 | 9 | 4 | 3 |
| Female | 31 | 12 | 5 | 1 |
| Transgender | 1 | 1 |  |  |
| Unknown |  |  |  |  |
| **Age** |  |  |  |  |
| 18-19 | 5 | 3 | 1 |  |
| 20-29 | 17 | 9 | 3 | 2 |
| 30-39 | 11 | 6 | 3 |  |
| 40-49 | 8 | 1 |  |  |
| 50-59 | 12 | 3 | 2 | 2 |
| 60-69 | 2 |  |  |  |
| Unknown |  |  |  |  |
| **Ethnicity** |  |  |  |  |
| Asian or Asian British | 2 | 1 |  |  |
| Mixed white and Asian | 2 |  |  |  |
| Black or Black British | 2 | 1 | 2 | 1 |
| Not stated/prefer not to say | 19 | 10 | 1 |  |
| White British/white other | 30 | 10 | 6 | 3 |

PharmOutcomes® data shows most PrEP consultations were initiated by pharmacists when community members attended the pharmacy for other health related services (n=25) including pharmacy opioid substitution therapy (e.g., methadone, n=11) or for emergency contraception (n=14). The number of PrEP consultations increased over time, particularly community member-initiated pharmacy PrEP consultations (n=14) (see figure 2).

**Figure 2.**
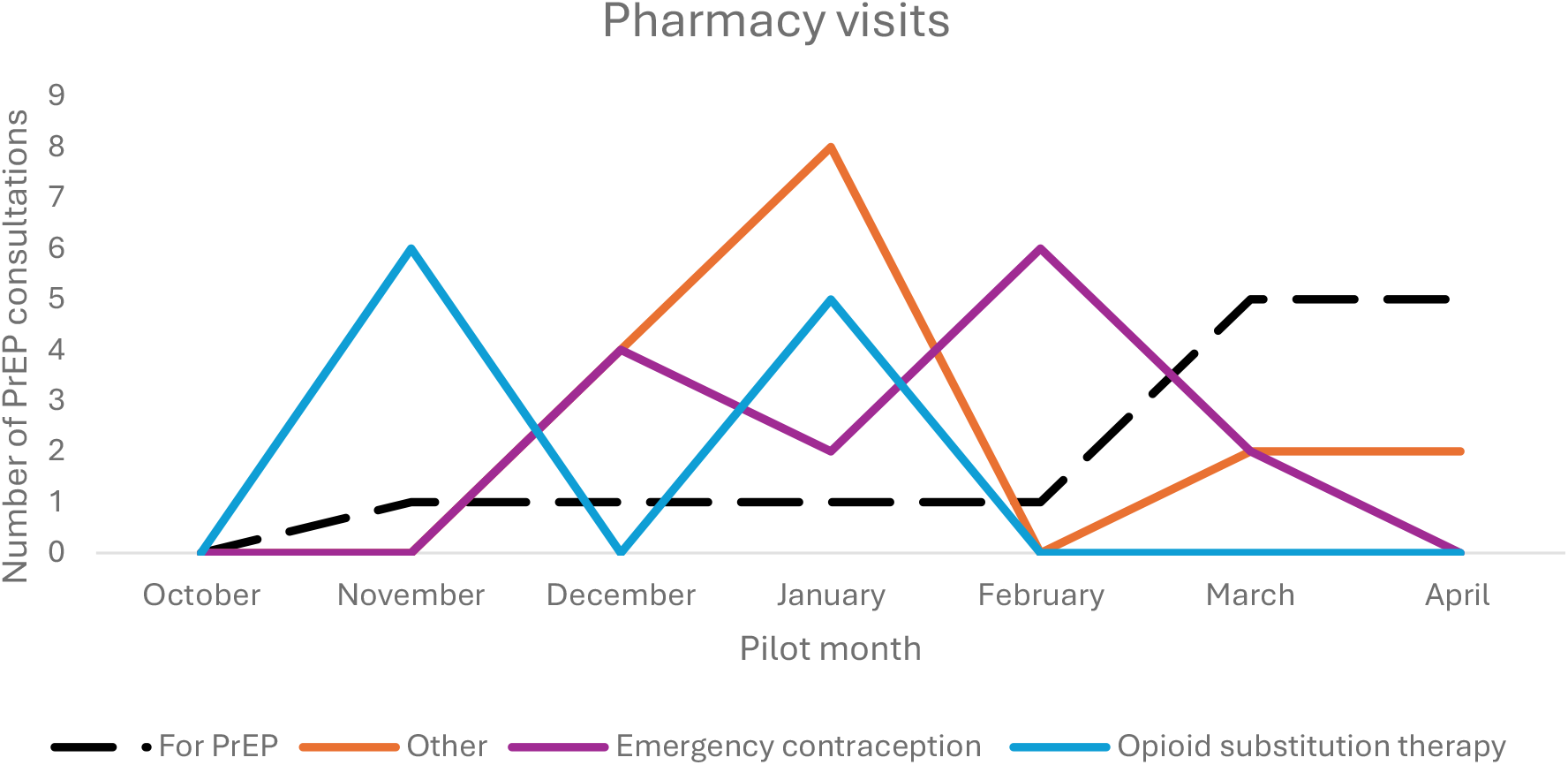
Number and reason for pharmacy visit for PrEP pharmacy consultation across the six-month pilot.

Of the 55 individuals who had a PrEP consultation, 28 (51%) were interested in PrEP and 22 (88%) of those interested took a testing kit home and were referred to the sexual health clinic for PrEP. Individuals not taking a test kit predominantly reported that they would like more time to think about PrEP.

Of those referred to the clinic, nine returned their test kit. Of those returning their test kit, one was uncontactable, three declined PrEP and four were prescribed PrEP (see Table 1). Of those prescribed PrEP, three were already known to the sexual health clinic due to having previously ordered postal testing kits (n=2) or having been seen in person at the clinic (n=1). One individual was new to the service.

#### c. Qualitative evaluation

Seven community members participated in an interview and five pharmacists participated in two interviews during month 3 and 6 of the pilot interview. Community members were women (n=4) and men (n=3) aged between 24 and 54 years who identified as being of white (n=5) and Black (n=2) ethnicity, bisexual (n=3), gay (n=1) and heterosexual (n=3). Three were prescribed PrEP and four declined a PrEP referral.

Analysis led to the development of the key emergent themes outlined below, reflecting the perspectives of pharmacists and community members. Illustrative quotations (Table 2) are followed by participant identifier (community member [CM], pharmacist [P]), number and interview time (T1/T2) for pharmacists).

**Table 2.**
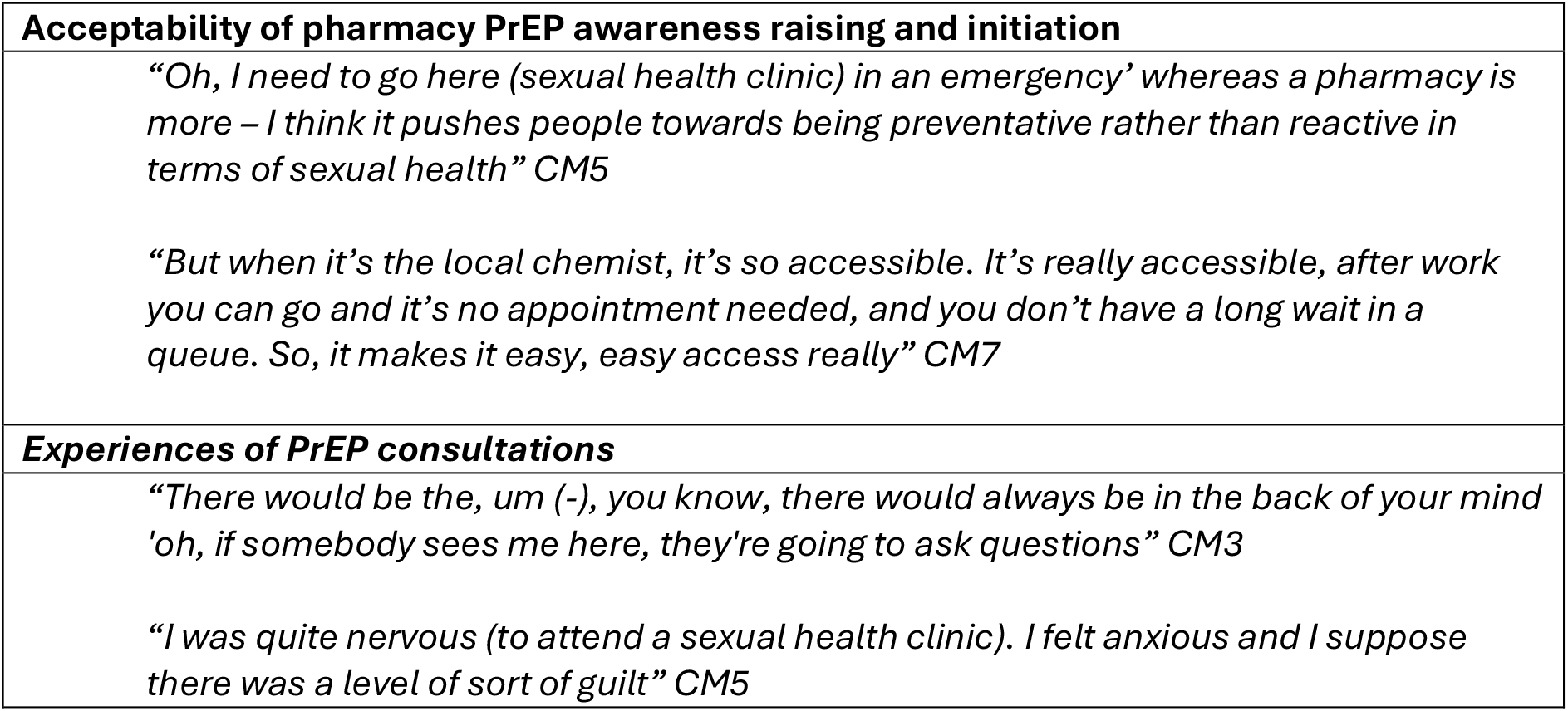
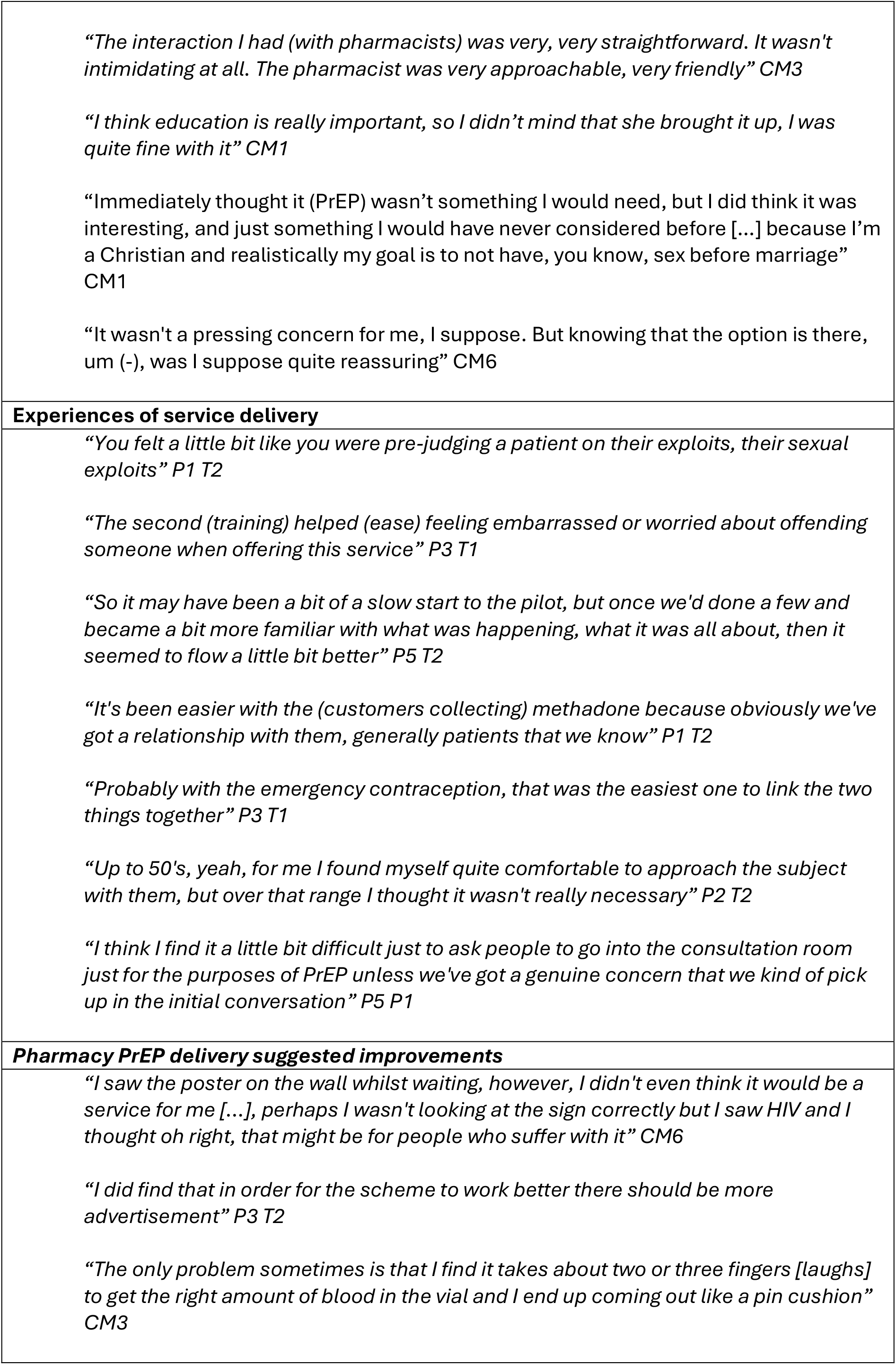
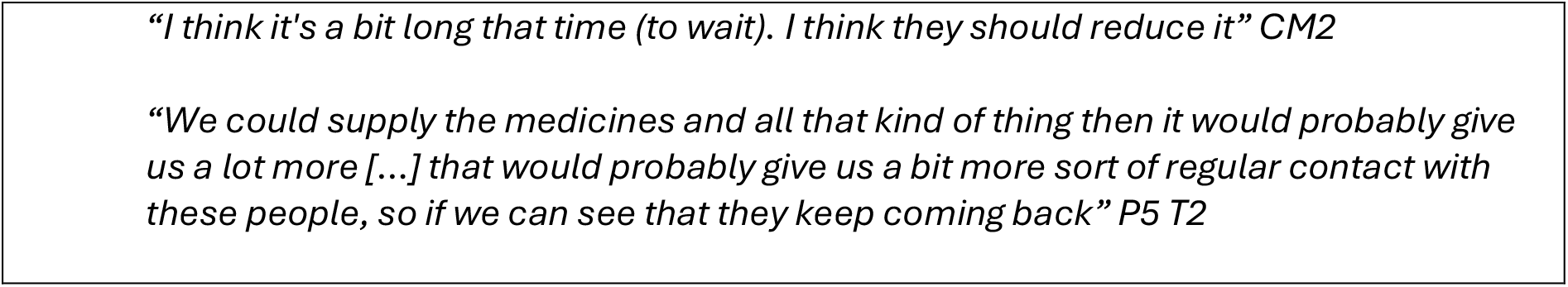
Illustrative quotes from the interviews for the four themes.

##### Acceptability of pharmacy PrEP awareness raising and initiation

Pharmacists and community members viewed pharmacy PrEP initiation positively, reporting pharmacies to be an important, beneficial option outside of sexual health clinics. They highlighted pharmacies to be useful points of contact for preventative rather than reactive service provision, their accessibility and anonymity, meant they had potential to help raise awareness of PrEP and reach individuals not currently aware of or accessing PrEP.

##### Experiences of PrEP consultations

Most community members who were interviewed had a history of online STI and/or HIV testing, but only two had previously attended a sexual health clinic in person. Previous decisions to use online testing kits were reported to be influenced by their convenience, accessibility and privacy. Those who had previously visited sexual health clinics reported their visits to be accompanied by feelings of shame, vulnerability, stigma, anxiety and a concern about being seen by someone they know.

Community members having pharmacy PrEP consultations reported having had positive experiences and feeling comfortable discussing PrEP with the pharmacist. Individuals who were introduced to PrEP while visiting the pharmacy for other services appreciated being informed about the medication, especially as they reported having never heard of it before their visit to the pharmacy. Those who actively sought PrEP from the pharmacy described the process as accessible and relatively straightforward.

Of those who were unaware of PrEP, one individual initiated PrEP as a result of being made aware of it during a visit to the pharmacy for emergency contraception. Of those declining PrEP, the primary reason why was because it was not seen to be personally relevant because of their faith and intending to abstain from sex before marriage, being in a stable relationship or having a regular partner and/or preferring behaviour modification over taking medication.

##### Experiences of service delivery

Pharmacists delivering the PrEP consultations reported experiencing some initial challenges implementing the pilot. These challenges primarily related to feelings of embarrassment, unease or concern about how best to bring up the conversation of PrEP with customers due to a lack of customers actively approaching them for PrEP. This was particularly pertinent at the beginning of the pilot because of concerns about causing offence or being perceived as judgemental. Confidence increased after the second in person training session and with experience.

The pharmacist-initiated PrEP consultations were more commonly undertaken with regular customers with pre-existing relationships, particularly those collecting methadone prescriptions. Pharmacists also found it easier to introduce the topic of PrEP in consultations that were already related to sexual health e.g. emergency contraception.

In other cases, pharmacists made quick assessments based on timing, context, and customer characteristics to determine whether initiating a PrEP discussion was judged appropriate. They often chose not to initiate these conversations with individuals in long-term relationships or those perceived to be older, as such customers were considered less likely to benefit from PrEP.

The practicalities of initiating a consultation also played a role. Pharmacists reported that PrEP discussions were more feasible during quieter periods, outside peak times like the winter months, when workload pressures were lower. Turning an over-the-counter inquiry into a PrEP consultation could be challenging as it required tactfully moving the customer into a private consultation room.

##### Pharmacy PrEP delivery suggested improvements

Although the community members and pharmacists perceived PrEP initiation via pharmacies positively, they highlighted several key areas for improvement to ensure the feasibility and effectiveness of the service. Pharmacists highlighted the low number of customers actively seeking PrEP. In contrast community members highlighted their lack of awareness of PrEP and its availability though pharmacies. Most community members reported not seeing the PrEP awareness raising material distributed during the pilot online, in community or in pharmacies. Those who did, often overlooked them, perceiving them to not be personally relevant.

To address this gap, both community members and pharmacists recommended more targeted and widespread campaigns, via social media and television, more prominent posters, and including leaflets in prescription bags. These efforts were seen as crucial not only to encourage individuals to initiate PrEP consultations but also to increase the perceived personal relevance of PrEP and help reduce HIV and sexual health-related stigma.

There were also concerns about the complexity of the screening test kits, particularly regarding complexity of instructions for non-English speaking individuals, completing the blood test at home and the time delay between completion of test kit and receiving the PrEP prescription.

The ability for pharmacists to hold PrEP stock themselves and provide their customers with immediate access would streamline felt the PrEP initiation pathway and strengthen ongoing relationships with repeat prescription customers.

## Discussion

This is the first UK pilot to explore the acceptability and feasibility of pharmacies raising awareness of and initiating oral PrEP with a particular focus on underserved community members. This novel pilot demonstrates pharmacy PrEP delivery to be acceptable and feasible, overall, successfully raising awareness of PrEP. Notably, this pilot reached a high percentage of women and PWID, groups that are shown to be less likely to be aware of and engage with traditional PrEP services. Data supports pharmacy based PrEP delivery, but highlights the need to build pharmacist confidence, raise community awareness and reduce structural and logistical barriers to allow direct pharmacy PrEP provision and maximise the reach and sustainability of pharmacy PrEP delivery.

UK community pharmacists already have an established role in sexual and reproductive health by providing emergency contraception, chlamydia screening, condom distribution and supplying oral contraception. The expansion of pharmacy clinical services has required pharmacists to undertake specialist training and establish new consultation routines driven by set protocols. The successful integration of these services demonstrates pharmacists’ capacity to acquire the skills and competencies needed to deliver sexual health services. This highlights their potential to support future expansion of PrEP delivery and extend access beyond traditional sexual health clinics [22, 23]. Nevertheless, despite the current pilot demonstrating acceptability of pharmacy PrEP delivery for pharmacists and community members, pharmacists did express initial concerns with initiating conversations about PrEP with their customers. This challenge is not unique to pharmacies but reflects early-stage difficulties associated with embedding new practices in routine care [13, 23]. However, as seen with other clinical services, pharmacists’ confidence, comfort and perceived capability improved over time with increased training and experience in PrEP delivery. Consistent with previous pharmacy PrEP delivery research, findings underscore the importance of accessible and high-quality pharmacy staff training and ongoing support to ensure they are equipped to deliver PrEP effectively [7, 15, 24].

Data on UK PrEP uptake shows the majority of PrEP users to be white GBMSM [5, 25]. Disproportionately low uptake has been observed among some community groups at elevated risk of acquiring HIV, such as trans individuals, women, young people, PWID and people of Black African or Black Caribbean heritage [26-30]. Previous research highlights a lack of awareness of PrEP as a potential barrier to PrEP initiation [7]. This pilot leveraged community pharmacies’ accessibility and provision of other existing sexual health, and clinical services, to engage community groups who may not otherwise be aware of or accessing PrEP. Results suggest that by expanding opportunities to engage individuals in discussions about PrEP, pharmacies could play a strategic role in improving awareness of PrEP for underserved groups. For example, a high proportion of the community members who received a PrEP consultation were aged ≤29 years, women and/or PWID and received a PrEP consultation while collecting emergency contraceptive or opioid replacement therapy. The majority interviewed were unaware of PrEP prior to their consultation. Embedding PrEP consultations into existing service specifications and subsequent consultations, such as emergency contraception and chlamydia testing, could help to routinise conversations, improving awareness of PrEP and support normalisation of its use beyond GBMSM communities. This could additionally help support broader system wide approaches for tackling HIV stigma. Routinising PrEP conversations may also help avoid any unconscious bias and selective offering of PrEP among pharmacists that may reinforce inequalities in service provision.

While community pharmacies could play a significant role in raising PrEP awareness and accessibility, structural and logistical barriers were evident, such as work capacity, complex referral pathways, pharmacists’ inability to perform the monitoring tests and stock and supply NHS PrEP directly. NHS England fund PrEP, and delivery is funded through the Public Health Grant to local authorities who commission sexual health services. Consequently, for the current pilot, a collaborative practice agreement between the five pharmacies and the local sexual health clinic was established to work barriers and enable pharmacy PrEP delivery without customers having to visit a sexual health clinic. The complexity and lack of timeliness of the pathway may have influenced completion of screening tests and PrEP uptake. For example, 32% of home test kits provided were returned, which is lower than the 51% return rate for home STI test kits obtained from vending machines and the 65% rate for local online STI testing services in BNSSG [33]. The lower return rate may underscore difficulties experienced when completing the extra blood tests at home required for PrEP initiation and the potential need to have in person support available for some [7]. Future research should explore the need for in person support, translated materials and the provision of pharmacy private spaces to facilitate completion of the screening tests, particularly for individuals with lower levels of English literacy, PWID who have no fixed abode and individuals who struggle to take their own blood.

Another possible reason for the low return rate may partly reflect individuals who took a test kit but did not return it before the pilot concluded.

Our results suggest that some community members may have reconsidered their decision to initiate PrEP following their pharmacy consultation and referral, or that referrals were lost within the system. For example, three individuals who returned their test kit subsequently declined PrEP, and one was uncontactable. Developing pathways that enable pharmacists to supply and manage PrEP autonomously may overcome these barriers. The increasing numbers of independent prescribing pharmacists may reduce reliance on manual referral systems and streamline PrEP access through pharmacies. However, direct pharmacy PrEP provision also requires pharmacies to be able to stock publicly funded (NHS) PrEP. Without access to NHS PrEP, pharmacists would need to procure PrEP at high costs, which would be prohibitively expensive for local authorities (commissioners of public sexual health services), pharmacies and customers [7, 15, 24]. Providing an autonomous free and comprehensive service may become particularly important with the introduction of long-acting injectable PrEP (i.e., cabotegravir and lenacapavir) alongside increasing pressures within sexual health services.

Since cabotegravir requires administration every eight weeks, and lenacapavir every six months, community pharmacies could serve as additional points of contact for initiation and continuation of care, offering significant support for sexual health services [36]. Future research should be conducted to determine the acceptability and feasibility of delivering long-acting injectable PrEP from community pharmacies.

## Strengths and limitations

The development of the pilot utilised robust PBA methods, integrating insights from academic literature, qualitative interviews, PPI and behavioural science theory to ensure its accessibility, appropriateness and relevance for underserved community members. Interviews with pharmacists and community members provided important insights into the experiences of delivering and receiving pharmacy PrEP consultations and suggestions for optimisation and sustainability. However, findings are based on data from pharmacists and community members in one geographical location and should be interpreted considering this. Future research should investigate community pharmacy PrEP delivery on a larger scale and via other healthcare services such as primary care [13].

## Conclusion

The pilot highlights that community pharmacy is not only a feasible but also a potentially transformative setting for PrEP delivery. With appropriate training, support, policy alignment and commissioning, pharmacy based PrEP delivery could play a critical role in reducing HIV transmission and address inequalities in sexual health.

## Data availability statement

Data are available in a public, open access repository at data-bris

## Ethical approval

Ethical approval for this study was awarded by the National Health Service, Health Research Authority, Research Ethics Committee (345513). All participants gave informed consent to participate in the study before taking part.

## Authors contributions

CH, HF, JK, SD, JS, CS, JC, LH, SC, JS, RHS, NS, OBD, ET and JH contributed to the research planning, CH to data collection and CH and HF to data analysis. CH drafted all versions of the manuscript, and all authors contributed to and approved the final version of the publication.

## Data availability

The data underlying this article are available at data-bris.ac.uk

## References

1. McCormack, S., et al., Pre-exposure prophylaxis to prevent the acquisition of HIV-1 infection (PROUD): effectiveness results from the pilot phase of a pragmatic open-label randomised trial. Lancet, 2016. 387(10013): p. 53–60.

2. Brady, M., et al., BHIVA/BASHH guidelines on the use of HIV pre-exposure prophylaxis (PrEP) 2018. HIV medicine, 2019. 20(S2): p. S2–S80.

3. UK health Security Agency. HIV testng, PrEP, new HIV diagnoses and care outcomes for people accessing HIV services: 2024 Report. 2024.

4. Shah AM.N.,, Ratna N, et al.,HIV testing,prep,new hiv diagnoses and care outcomes for people accessing hiv services: 2023 report. In: The annual official statistics data release (data to end of December 2022) 2023.

5. Coukan,F.,et al.,Barriers and facilitators to HIV pre-exposure prophylaxis (prep) in specialist sexual health services in the United kingdom: a systematic review using the prep care continuum. HIV medicine,2023. 24(8): p. 893–913.

6. Nydegger,L.A.,J. Dickson-Gomez,and T.K. Ko,Structural and syndemic barriers to PrEP adoption among Black women at high risk for HIV: a qualitative exploration. Culture,Health & Sexuality,2021. 23(5): p. 659–673.

7. Harrison,C.,et al.,Qualitative exploration of the barriers and facilitators to community pharmacy PrEP delivery for UK pharmacists and underserved community members using the COM-B model of behaviour change. Sex Transm Infect,2024.

8. Calabrese,S.K.,Understanding,contextualizing,and addressing PrEP stigma to enhance PrEP implementation. Current Hiv/Aids Reports,2020. 17: p. 579–588.

9. Sun,Z.,et al.,Increasing awareness of HIV pre-exposure prophylaxis (PrEP) and willingness to use HIV PrEP among men who have sex with men: a systematic review and meta-analysis of global data. Journal of the International AIDS Society,2022. 25(3): p. e25883.

10. Nagai,H.,et al.,HIV pre-exposure prophylaxis uptake among high-risk population in sub-Saharan Africa: A systematic review and meta-analysis. AIDS Patient Care and STDs,2024. 38(2): p. 70–81.

11. Department of Health and Social Care.,Towards Zero: the HIV action plan for England - 2022 to 2025. 2021.

12. Clutterbuck,D.,M. Brady,and A. Rodger,Driving progress towards PrEP equity: Key changes in the 2025 BASHH/BHIVA UK PrEP guidelines. 2025,Wiley Online Library.

13. Scott,A.,et al.,Developing the PATH-GP (Prevention and Testing for HIV in General Practice) intervention: a Person-Based Approach intervention development study to increase HIV testing and PrEP access. medRxiv,2025: p. 2025.03. 03.25323009.

14. BASHH/BHIVA.,Guidelines on the use of HIV pre-exposure prophylaxis (PrEP) 2025. 2025.

15. Harrison,C.,et al.,Facilitators and barriers to community pharmacy PrEP delivery: a scoping review. J Int AIDS Soc,2024. 27(3): p. e26232.

16. Zhao,A.,et al.,Pharmacy-Based Interventions to Increase Use of HIV Pre-exposure Prophylaxis in the United States: A Scoping Review. AIDS and Behavior,2022. 26(5): p. 1377–1392.

17. Department of Health and Social Care.,owards Zero – An action plan towards ending HIV transmission,AIDS and HIV-related deaths in England 2022 to 2025. GOV.UK. 2021.

18. Yardley,L.,et al.,The Person-Based Approach to Intervention Development: Application to Digital Health-Related Behavior Change Interventions. J Med Internet Res,2015. 17(1): p. e30.

19. Michie,S.,M.M. van Stralen,and R. West,The behaviour change wheel: A new method for characterising and designing behaviour change interventions. Implementation Science,2011. 6(1): p. 42.

20. Terrance HIggins Trust. PrEP training for pharmacies. Available from: https://www.tht.org.uk/about-us/what-we-do/training/prep-training-pharmacies.

21. Braun,V. and V. Clarke,Using thematic analysis in psychology. Qualitative research in psychology,2006. 3(2): p. 77–101.

22. Parsons,J.,et al.,Evaluation of a community pharmacy delivered oral contraception service. Journal of Family Planning and Reproductive Health Care,2013. 39(2): p. 97.

23. Chirewa,B. and A. Wakhisi,Emergency hormonal contraceptive service provision via community pharmacies in the UK: a systematic review of pharmacists’ and young women’s views,perspectives and experiences. Perspectives in Public Health,2020. 140(2): p. 108–116.

24. Alter,M.,et al.,Investigating facilitators and barriers to the routine provision of HIV PrEP in community pharmacies in London. BMC Health Services Research,2025. 25(1): p. 312.

25. Public Health Scotland.,HIV in scotland. 2023. Available from: https://publichealthscotland.scot/publications/hiv-in-scotland/hiv-in-scotland-update-to-31-december-2023/

26. Dang,M.,et al.,Barriers and facilitators to HIV pre-exposure prophylaxis uptake,adherence,and persistence among transgender populations in the United States: a systematic review. AIDS Patient Care and STDs,2022. 36(6): p. 236–248.

27. Wilson,E.C.,et al.,Knowledge,indications and willingness to take pre-exposure prophylaxis among transwomen in San Francisco,2013. PLoS One,2015. 10(6): p. e0128971.

28. Wu,H.,et al.,Uptake of HIV preexposure prophylaxis among commercially insured persons—United States,2010–2014. Clinical Infectious Diseases,2017. 64(2): p. 144–149.

29. Wayal,S.,et al.,The role of PrEP in the UK: investigating the barriers and facilitators for PrEP implementation among Black African and White MSM in London. BMC Public Health,2019. 19(1): p. 1578.

30. Nakasone,S.E.,et al.,Risk perception,safer sex practices and PrEP enthusiasm: barriers and facilitators to oral HIV pre-exposure prophylaxis in Black African and Black Caribbean women in the UK. Sexually transmitted infections,2020. 96(5): p. 349–354.

31. Estcourt,C.S.,et al.,HIV PrEP: raise awareness in all groups who could benefit and provide for both on and offline access. BMJ,2022. 378: p. o2133.

32. Kudrati,S.Z.,K. Hayashi,and T. Taggart,Social Media & PrEP: A Systematic Review of Social Media Campaigns to Increase PrEP Awareness & Uptake Among Young Black and Latinx MSM and Women. AIDS and Behavior,2021. 25(12): p. 4225–4234.

33. Gobin,M.,et al.,Acceptability of digital vending machines to access STI and HIV tests in two UK cities. Sexually Transmitted Infections,2024. 100(2): p. 91–97.

34. Velloza,J.,et al.,A Review of Implementation Strategies to Enhance PrEP Delivery for People Experiencing Housing Insecurity: Advancing a Multifaceted High-Touch,Low-Barrier Approach. Curr HIV/AIDS Rep,2024. 22(1): p. 4.

